# Comparison of two commercial molecular tests and a laboratory-developed modification of the CDC 2019-nCOV RT-PCR assay for the qualitative detection of SARS-CoV-2 from upper respiratory tract specimens

**DOI:** 10.1101/2020.05.02.20088740

**Authors:** Nicholas M. Moore, Haiying Li, Debra Schejbal, Jennifer Lindsley, Mary K. Hayden

## Abstract

We compared the ability of 2 commercial molecular amplification assays [*Real*Time SARS-CoV-2 on the *m*2000 (Abbott) and ID NOW™ COVID-19 (Abbott)] and a laboratory-developed test [modified CDC 2019-nCoV RT-PCR assay with RNA extraction by eMag^®^ (bioMérieux) and amplification on QuantStudio™ 6 or ABI 7500 Real-Time PCR System (Life Technologies)] to detect SARS-CoV-2 RNA in upper respiratory tract specimens. Discrepant results were adjudicated by medical record review. 200 nasopharyngeal swab specimens in viral transport medium (VTM) were collected from symptomatic patients between March 27 and April 9, 2020. Results were concordant for 167 specimens (83.5% overall agreement), including 94 positive and 73 negative specimens. The *Real*Time SARS-CoV-2 assay on the m2000 yielded 33 additional positive results, 25 of which were also positive by the modified CDC assay but not by the ID NOW™ COVID-19 assay. In a follow-up evaluation, 97 patients for whom a dry nasal swab specimen yielded negative results by the ID NOW™ COVID-19 assay had a paired nasopharyngeal swab specimen collected in VTM and tested by the *Real*Time SARS-CoV-2 assay; SARS-CoV-2 RNA was detected in 13 (13.4%) of these specimens. Medical record review deemed all discrepant results to be true positives. The ID NOW™ COVID-19 test was the easiest to perform and provided a result in the shortest time: as soon as 5 minutes for positive and 13 minutes for negative result. The *Real*Time SARS-CoV-2 assay on the m2000 detected more cases of COVID-19 infection than the modified CDC assay or the ID NOW™ COVID-19 test.

## INTRODUCTION

In December 2019, a cluster of patients with pneumonia of unknown origin was linked to exposure to a wet market in Wuhan, Hubei Province, China (1). Very quickly, a novel betacoronavirus was isolated from a lower respiratory tract sample from one patient and the full genome of the virus was sequenced (2). This novel coronavirus, which was named SARS-CoV-2 for its genetic homology to SARS-CoV, spread rapidly across the globe (3–10). As of April 29, 2020, more than 3 million cases of SARS-CoV-2 infection had been identified worldwide, with over 200,000 deaths; approximately one-third of cases have been identified in the United States.

Laboratory testing plays a critical role in defining the disease characteristics and epidemiology of an emerging infectious pathogen such as SARS-CoV-2, and in controlling its spread. Early on, laboratory testing for SARS-CoV-2 in the U.S. was performed only at the Centers for Disease Control and Prevention (CDC) laboratories in Atlanta, GA using a reverse transcriptase polymerase chain reaction (RT-PCR) assay that was developed there (11). Subsequently, the CDC test was to be implemented in all state public health laboratories, but roll out was slow due to technical problems. Following the declaration of a public health emergency, the US Food and Drug Administration (FDA) moved to allow *in vitro* diagnostic assays under an Emergency Use Authorization (EUA) in an attempt to expedite test development by commercial and clinical laboratories. The majority of assays approved through EUA are nucleic acid amplification tests that target conserved regions of the SARS-CoV-2 genome. Abbott Molecular received authorization for the *Real*Time SARS-CoV-2 assay to be performed on the m2000 real-time platform on March 18, 2020 (12). The ID NOW™ COVID-19 assay was granted approval under the EUA on March 27, 2020. *In vitro* diagnostic device (IVD) assays with EUA status from commercial manufacturers do not undergo usual FDA review under the *De Novo* request or the 510(k) premarket notification; as such, limited data comparing these assays are available.

In this study, we compared the performance of two commercial EUA IVD assays and a laboratory developed test that is a modification of the CDC RT-PCR assay for the qualitative detection of SARS-CoV-2 RNA directly from upper respiratory tract specimens.

## MATERIALS AND METHODS

### Clinical samples

For the initial evaluation of the three test systems, we collected nasopharyngeal swab specimens in 3mL M4-RT viral transport medium (VTM) (Remel, Lenexa, KS) from symptomatic (fever or cough or shortness of breath) adult and pediatric outpatients, emergency department (ED) patients, and inpatients at Rush University Medical Center (RUMC) or Rush Oak Park Hospital (ROPH); both hospitals are in metropolitan Chicago, IL. Specimens were collected between March 27 and April 9, 2020, and tested within 72 hours of collection; specimens were held refrigerated at 4°C if all testing could not be completed on the same day.

In a separate follow up evaluation, symptomatic patients who had a negative result on a dry nasal swab that was tested at the point of care by the ID NOW™ COVID-19 system also had a paired nasopharyngeal swab sample collected and transported to the on-site clinical microbiology laboratory for testing by *Real*Time SARS-CoV-2 on the *m*2000.

Age, sex, and location of swab collection were extracted from the electronic medical record (EMR) for all patients. The study was reviewed and given expedited approval by the RUMC institutional review board, with a waiver of written informed consent.

### Modified CDC assay

We validated and implemented a modification of the CDC 2019-nCoV assay (11) for clinical use in our laboratory; this was the first SARS-CoV-2 RT-PCR assay we adopted during the COVID-19 pandemic. This assay targets two regions of the nucleocapsid (N) gene of the SARS-CoV-2 genome. The human RNase P (RP) gene target is included and used as specimen extraction and amplification control.

Nucleic acids were purified and extracted using the eMAG® automated nucleic acid sample extraction system (bioMérieux, Marcy l’etoile, France). Briefly, total nucleic acids were extracted from VTM using a programmed input sample volume of 200μL into 2000μL of easyMAG® lysis buffer with the Specific B protocol to which a final eluted volume of purified nucleic acids was 50μL. We utilized the TaqPath™ 1-step RT-qPCR master mix (Life Technologies, Frederick, MD) and the 2019-nCoV CDC EUA kit (Integrated DNA Technologies, Coralville, IA) for target detection. Amplification and real-time detection was performed on the ABI 7500 Real-Time PCR System (Life Technologies) with software version 2.3, or on the QuantStudio™ 6 Flex Real-time PCR system (Life Technologies) using software version 1.4. The total sample volume per reaction was 15μL of master mix, combined primer/probe mix, and nuclease free water and 5μL of eluted sample. Assay run parameters were as described in the CDC protocol (11). Samples that gave a cycle threshold (C_t_) value <40 for both N1 and N2 targets were considered positive. Samples negative for both N1 and N2 targets had to have a positive amplification curve for the RP gene to be considered a valid negative result. Samples that gave a C_t_ value <40 for either N1 or N2 targets were considered inconclusive and repeat testing was performed per CDC protocol. If results were still inconclusive after repeat testing, a result of inconclusive was reported. Validation results for the laboratory-modified CDC assay are not shown.

### Abbott Molecular SARS-CoV-2 assay

Next, we verified the *Real*Time SARS-CoV-2 assay (Abbott Molecular, Des Plaines, IL), which is a qualitative real-time assay performed on the Abbott m2000 platform (12). The system includes the m2000sp instrument with automated extraction of nucleic acids using the DNA (total nucleic acid) sample preparation kit in batches of up to 96 samples. The *Real*Time SARS-CoV-2 assay utilizes two real-time detection probes: one probe combined for the N and RNA-dependent RNA polymerase (RdRP) genes, and a second probe for the internal control to assess overall performance, including nucleic acid extraction and possible PCR inhibition. Nasopharyngeal swab samples were heat inactivated at 56°C for 35±5 minutes prior to testing. Automated extraction was performed using a sample input volume of 500μL VTM, followed by automated addition of amplification pack reagents and extracts (40μL volume used for PCR amplification and detection). Two controls (one positive and one negative) provided by the manufacturer were included with each run of patient samples. Amplification curves were interpreted by the *m*2000rt system and reported as detected or not detected.

In our initial verification of the *Real*Time SARS-CoV-2 assay, we tested 25 nasopharyngeal swab samples in which SARS-CoV-2 RNA gene sequences had been detected by the laboratory-modified CDC assay, and 30 samples in which SARS-CoV-2 RNA samples were not detected. There was 100% positive and negative agreement between results of the 2 assays [median C_t_ values on the modified CDC assay for positive samples, 25.93 (IQR, 20.3 - 28.87) for N1 and 24.6 (IQR, 19.4 - 28.35) for N2].

### ID NOW™ COVID-19 assay

The third SARS-CoV-2 molecular assay introduced to our laboratory was the ID NOW™ COVID-19 (formerly Alere i), an isothermal nucleic acid amplification test for SARS-CoV-2 RNA that targets the RdRp gene (13). Following an initial 3 minute warm-up of the test system, 200μL of VTM was added to elution buffer in the sample base using the provided disposable transfer pipette, then mixed for 10 seconds with the pipette. Using the sample transfer device, the sample was transferred into the test cartridge, the lid was closed and the instrument automatically initialized the assay. The ID NOW does not report C_t_ values to the user. The instrument software interprets amplification data, and final results are reported on screen as positive, negative, or invalid.

### Estimation of RNA concentration in respiratory samples

We tested purified genomic RNA from a reference strain of SARS-CoV-2, isolate USA-WA1/2020 (catalog# NR-52285, lot# 70033320) (BEI Resources, Manassas, VA) to generate standard curves for the laboratory-modified CDC assay and the *Real*Time SARS-CoV-2 assay in order to estimate the concentration of SARS-CoV-2 genome equivalents in nasopharyngeal swab samples. We serially diluted the standard and tested the dilution series in triplicate. The ID NOW™ COVID-19 assay does not provide Ct values, so we could not generate a standard curve for this system.

### Data analysis

Because there is no reference standard for SARS-CoV-2 detection by RT-PCR, sensitivity and specificity of the assays could not be determined. Instead, we calculated overall, positive, and negative agreement for the 3 assays (http://www.medcalc.org). Box plots for comparing C_t_ values between groups were created using SPSS version 22 (IBM, Armonk, NY). Discordant results were adjudicated by medical record review (M.K.H.) to assess whether the patients’ clinical courses were consistent with COVID-19 infection.

## RESULTS

### Clinical overview

Specimens from 200 unique patients were included in this study. The first 94 samples tested for this study were collected consecutively. Subsequently, we enriched for positive samples by including all samples in which SARS-CoV-2 RNA was detected by the *Real*Time SARS-CoV-2 assay on the m2000 – our standard assay during the study period – and also the next negative sample after the positive sample. Mean age was 50 ± 17 years and 54% were women. Seventy-nine (40%) patients were hospitalized, 29 (36%) of whom were in an intensive care unit; 76 (38%) were cared for in an ambulatory location, including 55 (72%) who were seen in a designated COVID-19 screening clinic; and 45 (23%) were seen in an ED.

### Assay performance using nasopharyngeal swab samples in viral transport medium

There were 94 (47%) samples in which SARS-CoV-2 gene sequences were detected by all assays and 73 (36.5%) samples in which SARS-CoV-2 RNA was detected by none of the assays (Table 1). The median cycle number (C_n_) for positive samples by the *Real*Time SARS-CoV-2 assay was 15.34 (IQR, 11.27 - 18.13), or approximately 447 genome equivalents/μL (Figure 1). Overall agreement among the three assays was 83.5% (95% CI, 77.7% - 88.0%). Two-way positive and negative agreement between results is shown in Table 2. Positive agreement ranged from 75.2% to 100%, with the lowest agreement between the *Real*Time SARS-CoV-2 and the ID NOW™ COVID-19 assays. Negative agreement ranged from 92.4% (laboratory-modified CDC assay versus *Real*Time SARS-CoV-2 assay) and 100% (laboratory-modified CDC assay versus ID NOW™ COVID-19 assay, and ID NOW™ COVID-19 assay versus *Real*Time SARS-CoV-2 assay).

**Table 1.** Detection of SARS-CoV-2 RNA by laboratory-modified CDC 2019-nCOV RT-PCR assay, *Real*Time SARS-CoV-2 on the *m*2000, and ID NOW™ COVID-19^*^

| No. samples tested<br>(N=200) | Laboratory-modified CDC 2019-nCoV<br>RT-PCR | *RealTime* SARS-CoV-2 on the *m2000* | ID NOW COVID-19 |
| --- | --- | --- | --- |
| 94 | Detected | Detected | Detected |
| 73 | Not detected | Not detected | Not detected |
| 23 | Detected | Detected | Not detected |
| 2 | Detected | Detected | Invalid<sup>a</sup> |
| 6 | Not detected | Detected | Not detected |
| 2 | Inconclusive<sup>b</sup> | Detected | Not detected |

\*Categories with zero samples are not shown.
<sup>a</sup>Invalid defined as a sample that gave neither a positive nor a negative result.
<sup>b</sup>Inconclusive defined as a sample that gave a C<sub>t</sub> value <40 for either N1 or N2 targets.

**Figure 1.**
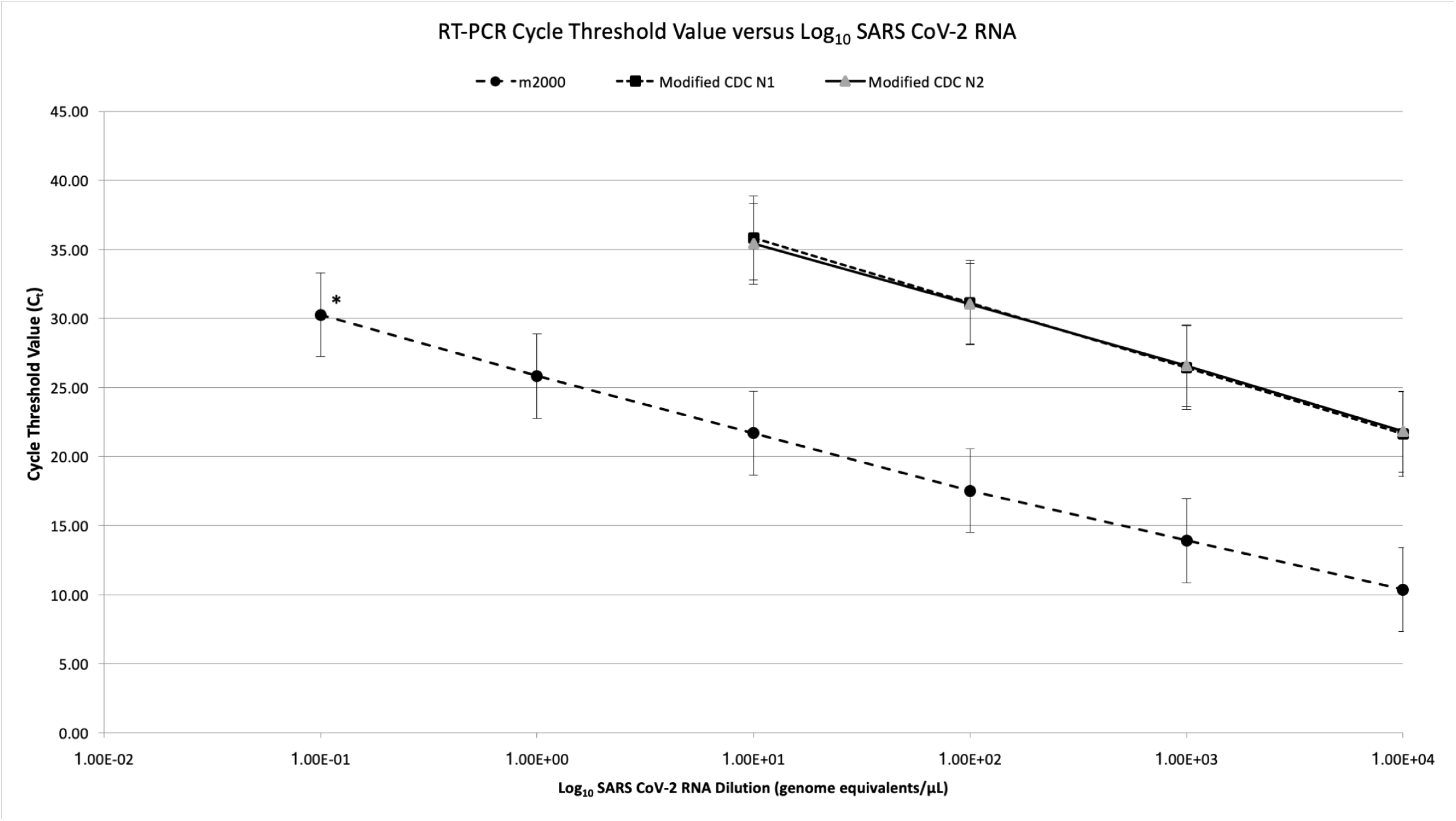
SARS-COV-2 Standard Curves for laboratory-modified CDC 2019-nCOV RT-PCR assay and *Real*Time SARS-CoV-2 on the *m*2000 assay. Values shown represent mean ± SEM of three independent replicates. Trend line equations: Laboratory-modified CDC assay (N1), y=-2.054ln(x) + 40.585, R2=1.0; laboratory modified CDC assay (N2), y=-1.966ln(x) + 40.022, R2 = 0.99; *Real*Time SARS-CoV-2 on the *m*2000 assay, y=-1.729ln(x) + 25.899, R^2^ =0.99. *Only 2 of 3 replicates amplified and are included in estimate.

**Table 2.** Performance agreement for detection of SARS-CoV-2 RNA by laboratory-modified CDC 2019-nCOV RT-PCR assay, *Real*Time SARS-CoV-2 on the *m*2000, and ID NOW™ COVID-19 (N=200 samples)

| Assay comparison (A/B) | Positive Percent Agreement (95% CI) |  | Assay comparison (A/B) | Negative Percent Agreement (95% CI) |
| --- | --- | --- | --- | --- |
| LDT+/m2000+ | 100 (96.9, 100) |  | LDT-/2000- | 92.4 (84.2, 97.2) |
| LDT+/ID NOW+ | 80.3 (71.9, 87.1) |  | LDT-/ID NOW- | 100 (95.4, 100) |
| m2000+/ID NOW+ | 75.2 (66.7, 82.5) |  | m2000-/ID NOW- | 100 (95.4, 100) |

Abbreviations: LDT, laboratory-modified CDC 2019-nCoV RT-PCR assay; m2000, *Rea/Time* SARS-CoV-2 on the *m2000*; ID NOW, ID NOW™ COVID- 19.

For the laboratory-modified CDC assay, SARS-CoV-2 target RNA sequences were detected in 119 (60%) samples. Six (3%) samples gave an initial inconclusive result. Upon repeat testing, 4 yielded valid results: 3 were detected and 1 was not detected. The remaining 2 (0.01%) samples repeated as inconclusive (only one of the two targets amplified in the specimen) (Table 1). The median C_t_ value for positive samples was 30.29 (IQR, 25.40 – 34.55) for N1 target and 30.20 (IQR, 25.12 – 34.55), which corresponds to an RNA concentration of approximately 150 genome equivalents/μL of sample (calculated using N1 standard curve) (Figure 1).

The *Real*Time SARS-CoV-2 on the *m*2000 assay yielded 127 (63.5%) positive results and no invalid results (Table 1). The median C_n_ value for positive samples was 17.27 (IQR, 13.27 - 21.40), which correlates to an RNA concentration of approximately 147 genome equivalents/μL of sample (Figure 1). The ID NOW™ COVID-19 assay yielded 94 (47%) positive results (Table 1). Five (0.03%) samples first gave invalid results; 3 resolved after repeat testing and the remaining 2 repeated as invalid.

### Analysis of discordant results

There were 33 (17%) samples that yielded discordant results across the three assays (Table 1). Eight discordant samples were not detected or gave inconclusive results by the laboratory-modified CDC assay but were detected by the *Real*Time SARS-CoV-2 assay. The median C_n_ value for these samples on the *Real*Time SARS-CoV-2 was 27.73 (IQR, 27.37 – 28.40), or approximately 0.34 genome equivalents/μL (Figure 2). Thirty-three samples (including 2 invalid samples) were not detected by the ID NOW™ COVID-19 but were detected by the *Real*Time SARS-CoV-2 assay; 25 of these were also detected by the laboratory-modified CDC assay. The median C_n_ values for these samples on the *Real*Time SARS-CoV-2 were 21.42 (IQR, 20.80 - 23.88), or approximately 13.3 genome equivalents/μL; differences among median values were statistically significant (Kruskal-Wallis p<.001) (Figure 2).

**Figure 2.**
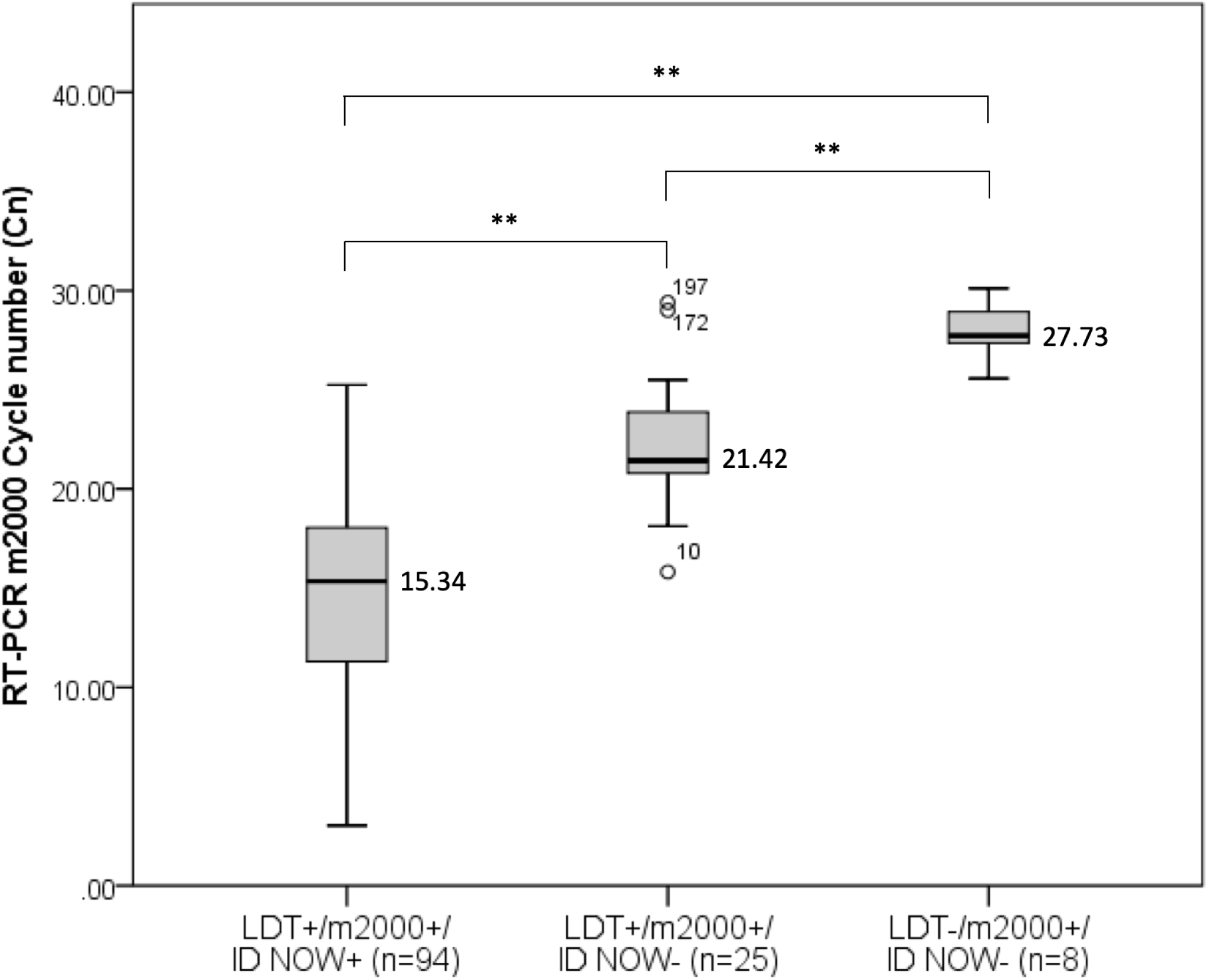
Comparison of Ct values among samples d 348 etected by each of the three assays, as measured by the *Real*Time SARS-CoV-2 on the *m*2000 standard curve. Median Ct value differences were statistically significant (**Kruskal-Wallis p<0.001 for all comparisons). Abbreviations: LDT, laboratory-modified CDC 2019-nCOV RT-PCR assay; *m*2000, *Real*Time SARS-CoV-2 on the *m*2000; ID NOW, ID NOW™ COVID-19.

Medical record review resolved all discrepant results in favor of the positive result (127 true positives). The *Real*Time SARS-CoV-2 assay detected significantly more cases of COVID-19 than either the laboratory-modified CDC assay [8 (4%) undetected cases, SD 0.014, 95% CI, 0.013 – 0.067] or the ID NOW™ COVID-19 assay [33 (16.5%) undetected cases, SD 0.026, 95% CI, 0.11 – 0.22]; the difference in detection between the laboratory-modified CDC assay and the ID NOW™ COVID-19 assay was also significant.

### Assessment of dry nasal swabs tested by the ID NOW™ COVID-19 assay

To analyze the performance of the ID NOW™ COVID-19 assay further, we tested its performance using dry nasal swab samples and compared results to those from paired nasopharyngeal samples tested by the *Real*Time SARS-CoV-2 assay. Ninety-seven patients were included in this evaluation. Mean age of patients was 59 ±17 years, and 48 (48%) were women. Thirteen (13.4%) paired nasopharyngeal samples yielded positive results by the *Real*Time SARS-CoV-2 assay. C_n_ values of these samples ranged from 9.2-29.2. SARS-CoV-2 RNA was not detected in the remaining 84 (86.6%) samples.

### Workflow analysis

Differences among key aspects of workflow for each of the three assays and platforms are summarized in Table 3. We estimate that we can batch test and report results for 58 samples by the laboratory-modified CDC assay, and 94 samples by the *Real*Time SARS-CoV-2 assay, in an 8-hour shift. The ID NOW™ COVID-19 assay, which was developed for point of care, requires the least hands-on time and provides the fastest results. However, throughput is limited (1 sample/instrument/5-15 minutes).

**Table 3.** Workflow analysis comparing laboratory-modified CDC 2019-nCQV RT-PCR, Abbott *m2000* SARS-CoV-2, and ID NOW™ COVID-19 assays

| Parameter<sup>a</sup> | Laboratory-modified CDC<br>2019-nCoV RT-PCR | *Real*Time SARS-CoV-2 on<br>the *m2000* | ID NOW™ COVID-19 |
| --- | --- | --- | --- |
| Off-board lysis | Yes | No | No |
| Specimen processing & set up | 1.75 | 1.00 | 0.03 |
| Instrument extraction time | 1.30 | 4.0 | 0.05 |
| Amplification & real-time detection | 1.25 | 2.25 | 0.22 |
| Manual interpretation and result entry | 0.5 | 0.75 | .02 |
| Total time to result<sup>b</sup> | 4.8 | 8.0 | 0.27 |
| No. samples processed in 8-hour shift per<br>instrument | 58<sup>c</sup> | 94<sup>d</sup> | 32<sup>e</sup> |

<sup>a</sup>Times are per batch for the in-house LDT and Abbott *m2000* assays, and per sample for the ID NOW.
<sup>b</sup>Time from sample processing through result reporting
<sup>c</sup>A maximum of 58 clinical samples, not including external positive control, negative control, negative template control, RNase P control.
<sup>d</sup>A maximum of 94 clinical samples.
<sup>e</sup>Assumes continuous processing of 32 clinical samples, all with negative results.

## DISCUSSION

Rapid, accurate detection of COVID-19 is essential to ensure speedy and appropriate patient management, outbreak containment, and to better understand the global epidemiology of the virus. Laboratory testing to date has relied primarily on the amplification and detection of viral gene sequences in upper respiratory tract specimens. As new test kits are made available through the EUA pathway, laboratories are confronted with the dilemma of deciding which test or platform to adopt for SARS-CoV-2 detection. Additionally, laboratory directors are faced with numerous questions from clinicians regarding performance characteristics of the tests. Responding to these questions is difficult, since EUA requires only limited test validation (14); assays approved under EUA have not been evaluated in clinical trials, and robust performance data from real world assessments are lacking.

Results of the current study help to fill this knowledge gap. We found significant differences in detection of SARS-CoV-2 viral sequences among the *Real*Time SARS-CoV-2 assay on the *m*2000, the laboratory-modified CDC assay, and the ID NOW™ COVID-19 assay. The *Real*Time SARS-CoV-2 assay on the *m*2000 detected the most cases, followed by the laboratory-modified CDC assay, and then the ID NOW™ COVID-19 assay. Discrepant results were observed almost exclusively in samples with higher C_t_ values, i.e., lower viral titer. These findings suggest differences in lower limit of detection of the assays. For the laboratory-modified CDC RT-PCR assay, this might be explained in part by smaller input sample volumes for extraction (200μL) and amplification (5μL), compared to 500μL extraction and 40μL amplification volumes in the *Real*Time SARS-CoV-2 assay on the m2000, *i.e*., there is more available target for amplification and detection in the *Real*Time SARS-CoV-2 assay on the *m*2000. Our results comparing the *Real*Time SARS-CoV-2 assay on the *m*2000 and the ID NOW™ COVID-19 assay are concordant with those of Harrington et al, who reported increased detection of SARS-CoV-2 RNA gene sequences by the *Real*Time SARS-CoV-2 assay compared to the ID NOW™ COVID-19 assay (15).

In order to eliminate confounding that could have been introduced by testing different sample types on different systems, we evaluated aliquots of the same nasopharyngeal swab/viral transport medium in all three assays. At the time of this study, nasopharyngeal swab specimens in viral transport medium were deemed acceptable sample types for the 3 assays that we assessed. Following the completion of our study, the manufacturer amended the package insert of the ID NOW™ COVID-19 assay to state that testing viral transport medium could lead to false-negative results. However, in our subsequent analysis of dry nasal swab samples tested by the ID NOW™ COVID-19 assay and paired nasopharyngeal swabs tested by the *Real*Time SARS-CoV-2 assay on the *m*2000, we continued to see more true positive results by the he *Real*Time SARS-CoV-2 assay on the *m*2000 assay, suggesting that false-negative results were not due entirely to dilution.

We observed differences in turnaround time, workflow, and throughput among the three tests. The *Real*Time SARS-CoV-2 assay on the *m*2000 had the longest runtime of the three assays: approximately 8 hours for one full run of 94 samples. Runtime of the laboratory-developed CDC assay was similar, but the throughput was less (58 samples in an 8-hour shift). The ID NOW™ COVID-19 assay was the easiest to perform and yielded the fastest results; positive results are generated in as few as 5 minutes, which is faster than any other test system available currently in the United States. Ease of use and speed are advantages in settings without laboratory expertise, or when rapid results are needed. The assay platform is small and can be utilized at near-patient settings, thereby increasing the overall testing capacity for SARS-CoV-2 within healthcare facilities. Availability of different platforms provides beneficial flexibility to meet testing needs of different populations and different healthcare settings.

Our study has limitations. Because there is not a reference standard for SARS-CoV-2 infection, we were unable to calculate sensitivity or specificity of the assays. Instead, we calculated percent agreement, which is appropriate when a non-standard reference method is utilized to compare assay performance (16). We resolved discrepant results through review of patient medical records, which may have introduced bias, since concordant test results were not confirmed in the same way (17). We enriched for samples that were positive by the *Real*Time SARS-CoV-2 assay on the *m*2000, which may have biased in favor of this test. However, our inability to detect any samples that yielded a positive result by either of the other two assays under evaluation and a negative result by the *Real*Time SARS-CoV-2 assay in this study, or in our initial verification of the assay, suggests that the effect of bias is small. Not all testing was performed on the same day due to workflow and personnel limitations, although all testing was completed within 72 hours of sample collection. Storage of specimens at ambient room (22°C) or refrigerated (4°C) temperature has been shown to have little impact on detection of other RNA viruses by RT-PCR (18).

In conclusion, we found that The *Real*Time SARS-CoV-2 assay on the m2000 detected more cases of COVID-19 infection than the modified CDC assay or the ID NOW™ COVID-19 test. The ID NOW™ COVID-19 test provided fastest results, and the small footprint of the instrument and ease of use are advantages in settings without technical expertise. Both tests are welcome additions to the COVID-19 testing armamentarium, and increase nationwide testing capacity for COVID-19.

## Data Availability

Anonymized data is available upon reasonable request.

## Acknowledgements

The following reagent was obtained through BEI Resources, NIAID, NIH: Genomic RNA from SARS-Related Coronavirus 2, Isolate USA-WA1/2020, NR-52285.

